# Family-Assisted Severity of Illness Monitoring for Hospitalized Children in Low-resource settings – a two-arm interventional pilot study

**DOI:** 10.1101/2021.11.16.21266433

**Authors:** Amelie O. von Saint Andre-von Arnim, Rashmi K. Kumar, Jonna D. Clark, Benjamin S. Wilfond, Quynh-Uyen P. Nguyen, Daniel M. Mutonga, Jerry Zimmerman, Assaf P. Oron, Judd L. Walson

## Abstract

**Introduction:** Pediatric mortality remains unacceptably high in many low-resource settings, with inpatient deaths often associated with delayed recognition of clinical deterioration. The Family-Assisted Severe Febrile Illness ThERapy (FASTER) tool has been developed for caregivers to assist in monitoring their hospitalized children and alert clinicians. While utilization of the tool is feasible, the impact on outcomes in low-resource settings has not been studied.

**Methods:** Randomized controlled pilot study at Kenyatta National Hospital, Kenya. Children hospitalized with acute febrile illness with a caregiver at the bedside for 24 hours were enrolled. Caregivers were trained using the FASTER tool (monitors work of breathing, mental status, perfusion, producing color-coded flags to signal illness severity). The primary outcome was the frequency of clinician reassessments between intervention (FASTER) and control (standard care) arms. Secondary outcomes included survey assessments of clinician and caregiver experiences with the tool. The study was registered with ClinicalTrials.gov NCT03513861.

**Results:** 150 patient/caregiver pairs were enrolled, 139 included in the analysis, 74 in the intervention, 65 in the control arm. Patients’ median age was 0.9 (range 0.2-10) and 1.1 years (range 0.2-12) in intervention versus control arms. The most common diagnoses were pneumonia (80[58%]), meningitis (58[38%]) and malaria (34[24%]). 134(96%) caregivers were patients’ mothers. Clinician visits/hour increased with patients’ illness severity in both arms, but without difference in frequency between arms (point estimate for the difference -0.2%, p=0.99). Of the 16 deaths, 8 (four/arm) occurred within 2 days of enrollment. Forty clinicians were surveyed, 33(82%) reporting that FASTER could improve outcomes of very sick children in low-resource settings; 26(65%) rating caregivers as able to adequately capture patients’ severity of illness. Of 70 caregivers surveyed, 63(90%) reported that FASTER training was easy to understand; all(100%) agreed that the intervention would improve care of hospitalized children and help identify sick children in their community.

**Discussion:** Although we observed no difference in recorded frequency of clinician visits with FASTER monitoring, the tool was rated positively by caregivers and clinicians. Further research to refine implementation with additional input from all stakeholders might increase the effectiveness of FASTER in detecting and responding to clinical deterioration in low-resource settings.

## Introduction

Pediatric mortality in resource-poor settings continues to be high, with under-five mortality rates in Africa in 2020 at 76 per 1000 children or 1 in every 13 children(1). These deaths are often due to preventable and treatable conditions, including neonatal diseases, lower respiratory tract infections and diarrheal illnesses (2). Management of severe illness in low-resource settings is often conducted on general hospital wards under significant resource constraints, rather than in intensive care units. According to the World Health Organization (WHO), the African region experiences both the greatest burden of disease and the lowest density of health workers at 2.2 healthcare professionals per 1000 population(3). This health care worker shortage results in overburdened medical staff, overcrowded facilities and limitations in the inpatient monitoring (4-6), with worsening illness often under-recognized and associated with substantial mortality (7) (8).

Prediction models that enhance early identification of the sickest children are needed to guide timely referral and transport of patients, efficient allocation of resources, and counselling regarding anticipated clinical trajectories (9) (10). Empowering family members to assist with timely recognition of clinical deterioration in their hospitalized child may allow for expedited clinical response and improve health outcomes. A prospective, feasibility study at Kenyatta National Hospital (KNH) in Nairobi examined the adequacy of the simple 3-point FASTER bedside assessment tool (Figure 1) as a potential method for enlisting caregivers to identify and communicate patient deterioration and demonstrated that FASTER assessment by caregivers is feasible in low-resource settings(11). In addition, caregiver assessments correlated strongly with professional research team assessments, using established severity of illness systems (Bedside Pediatric Early Warning Score or PEWS(12)), and with fatalities within the first 48 hours of admission(11). In the current report, we examine the impact of caregiver assessments and signaling using the FASTER monitoring tool on frequency of clinician assessments and explore caregiver and clinician experiences with this intervention.

**Figure 1:**
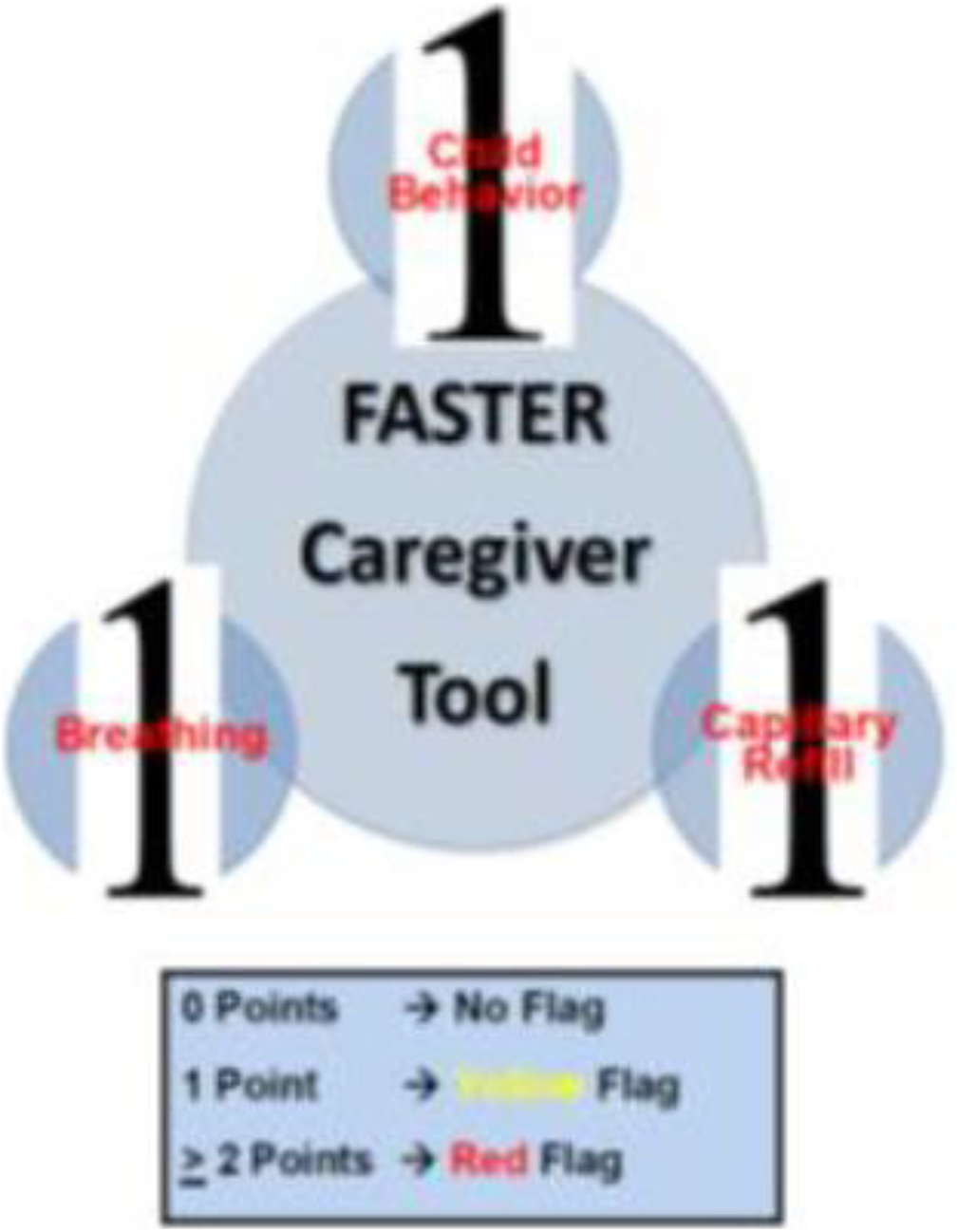
Caregiver FASTER Tool with 1 point given respectively for presence of chest retractions, capillary refill time > 3 sec and abnormal child behavior with patients responsive only to painful stimuli or non-responsive to pain.

## Materials and Methods

### Eligibility

Children aged 2 months to 12 years admitted to the KNH pediatric wards or “acute rooms” (ward rooms with higher nurse to patient ratios) within the previous 16 hours with severe febrile illness were eligible. Inclusion diagnoses included malaria, sepsis or septic shock, pneumonia, and meningitis or encephalitis. Patients were excluded if their primary diagnosis was related to major bleeding or hemorrhagic shock, severe trauma or burn, major surgery, known congenital heart disease, if an adult caregiver would not be consistently present for the entire 24 hour study period or if the caregiver was not proficient in English or Swahili.

### Intervention and Study Arms

Caregivers were enrolled 1:1 into intervention or control arms based on a weekly rotating schedule, until target sample size of 75 caregiver/patient pairs per arm was reached. Caregivers in the interventional arm received individualized education regarding family-assisted monitoring, which included video-based and hands-on training provided by a study nurse(11). Caregivers were taught to identify signs of clinical deterioration, namely: presence of chest retractions, capillary refill time > 3 sec, and an altered mental status (responsive only to painful stimuli or non-responsive).They were instructed to perform the clinical assessment every hour for 24 hours and display a color-coded severity of illness flag, with a red flag indicating high severity of illness (2 or more FASTER signs), a yellow flag for moderate severity of illness (one FASTER sign), and no flag for patients with zero FASTER signs (Figure 1). Control arm caregivers did not receive child clinical assessment training and did not participate in the FASTER clinical monitoring protocol. Caregivers in both arms, however, recorded the frequency of clinician visits to their child’s bedside during the first 24 hours post enrollment. Study team nurses performed the FASTER assessment on patients in both arms, 4 times during the 24-hour study period. Study team FASTER assessments were not shared with clinicians or caregivers given that correlation with validated severity of illness tools (Bedside PEWS) was not yet established at the time of this intervention. Caregivers in the intervention arm and all clinicians caring for children in both arms were surveyed about their experiences using the FASTER intervention.

Ethical approval was obtained from KNH/University of Nairobi and at Seattle Children’s Hospital. All caregivers provided written informed consent for participation in the study. The study was registered with ClinicalTrials.gov NCT03513861.

### Endpoints and Data Collection

Clinical data for the Bedside PEWS at and 24 hours post enrollment, study team FASTER scores, case fatality data and demographic information were collected and entered into a Research Electronic Data Capture (REDCap) form, hosted by the Institute for Translational Health Sciences at the University of Washington (12, 13). Caregivers recorded the FASTER flags raised and frequency of clinician visits on paper forms, which the study team later entered into REDCap.

### Sample Size and Statistical Analysis

A sample size (n=100) was calculated to enable detection of a relative increase of 50% in clinician visits to deteriorating patients (defined as FASTER “red-flag” assessment, i.e., 2 or more deterioration signs) on the intervention arm, compared with the control arm. These calculations assumed that 70% of caregiver assessments would be “red-flag”. However, the feasibility study revealed that only 10-15% of assessments were “red-flag(11). This lower than expected frequency indicated substantially reduced power for the main study. Therefore, the sample size was increased to n=150, the maximum sample feasible given budget and time constraints. This only allowed detection of a substantially larger effect than originally planned. The study was not powered to detect differences in mortality between arms. Power calculations and statistical analyses were carried out using R, versions 3.0 through 4.1 (R Foundation for Statistical Computing, Vienna).

The primary outcome was the effect of the intervention on the association between severity of illness and the number of clinician (nurse and physician) visits to the patient’s bedside. We used research-team FASTER assessments, available on both arms, as proxy for the child’s real-time condition severity. Since intervention-arm caregiver FASTER flags were very similar to research-team assessments (11), if caregiver flag affected provider behavior in the desired manner, then the difference in frequency of clinician bedside visits to patients with higher vs. lower research-team FASTER assessments would be greater on the intervention arm. This intervention effect was estimated in a regression model as an interaction between arm and FASTER assessment (dichotomized as red-flag vs. less severe), with the number of hourly visits being the response variable. We used Poisson regression with a random intercept for grouping by patient, adjusting for admission PEWS and age under 6 months.

In both arms, clinician visits were recorded hourly by caregivers. Missing-data patterns suggested that during late-night hours, most caregivers rested and did not record visits consistently; this time also coincides with lower clinician-visit frequency. We therefore performed the primary analysis on data collected between 06:00 to 22:00. During hours with no clinician visit, control-arm caregivers left the data entry form blank, whereas intervention-arm caregivers generally entered zeros for such hours [Supplemental Tabble 1]. To overcome this reporting difference, in the primary analysis we treated blank entries as zero. In sensitivity analysis, blank entries were excluded. In secondary analysis, we tested for a potential indirect clinical intervention impact by comparing the change in PEWS over 24 hours between arms, among surviving patients, using simple linear regression.

Survey data were collected from 40 health care providers and 70 caregivers to explore their perspectives regarding the benefits and challenges of the FASTER monitoring tool [Supplemental Material 2]. Using both open and closed ended questions, it assessed the overall clinician and caregiver experience with FASTER; challenges, general value and caregivers’ understanding of the tool. Research nurses recorded the verbal responses in either Swahili or English to survey questions from caregivers, as not all caregivers in the study were literate. Clinicians responded to survey questions in writing.

Qualitative responses to open ended survey questions by caregivers and health care providers were short and concise. One research team member categorized individual responses to each question into themes based on content (JC). Summaries of these data were created from the categorization of themes. Two additional research team members (BW, AV) reviewed the thematic categorization of survey responses and data summaries. Any differences in opinion were discussed and modified until consensus among the research team was achieved to improve reliability of the data summaries.

## Results

### Demographics of Study Population

Enrollment at KNH occurred between July and November 2017. Of the 150 caregiver/patient pairs enrolled, 139 were included in the analysis, 74 in the intervention arm and 65 in the control arm (Table 1). Two patients were excluded because they deteriorated and died so quickly that caregivers did not have time to record provider visits. Nine additional patients, all in the control arm, were excluded because no study-team FASTER assessments were performed. Among included patients, median age was 0.9 years (range 0.2-10) in the intervention arm and 1.1 years (range 0.2-12) in the control arm; with 38 (51%) and 23 (35%) female in intervention versus control arms. The most prevalent admission diagnoses in both arms were pneumonia (80 [58%]), meningitis (58 [38%]) and malaria (34 [24%]). Nearly all caregivers in both arms were patients’ mothers (134 [96%]), with the most common level of education being primary (47 [34%]) or secondary school (67 [48%]) (Table 1). Among included patients, 16 of 139 (12%) died in the hospital, nine of them on the intervention arm. Eight patients (four in each arm) died within two days of enrollment. Case-fatality rate did not vary by child age, however death within two days of enrollment was associated with age: 6 of 8 infant fatalities occurred within two days, compared with 2 of 4 deaths among those aged 12-23 months, and no fatalities among children 2 years or older; all 3 late deaths (> 1 one week) occurred in this age group (post-hoc Chi-Squared p=0.01). Admission PEWS was strongly associated with early death: 7 of 41 patients with bedside PEWS≥10 (17%) died within two days, compared with only 1 of 76 (1%) who had PEWS between 5 and 9, and 0 of 22 with PEWS<5 (p=0.003).

**Table 1.** Demographics FASTER invention and control arm for patient/caregiver pairs

| Patient/Caregiver Characteristic | Control<br>n (%) | Intervention<br>n (%) | Sum<br>n (%) |
| --- | --- | --- | --- |
| Patient number | 65 | 74 | 139 |
| Patient Median Age in years (range) | 1.1 (0.2-12) | 0.9 (0.2-10) |  |
| Sex |  |  |  |
| Male | 42 (65) | 36 (49) | 78 (56) |
| Female | 23 (35) | 38 (51) | 61 (44) |
| Caregiver |  |  |  |
| Mother | 62 (95) | 72 (97) | 134 (96) |
| Father | 1 (2) | 0 (0) | 1 (1) |
| Grandparent | 0 (0) | 1 (1) | 1 (1) |
| Aunt/Uncle | 2 (3) | 1 (1) | 3 (2) |
| Language |  |  |  |
| Swahili | 36 (55) | 48 (65) | 84 (60) |
| English | 29 (45) | 26 (35) | 55 (40) |
| Caregiver Highest Level of Education |  |  |  |
| Primary | 26 (40) | 21 (28) | 47 (34) |
| Secondary | 25 (38) | 42 (57) | 67 (48) |
| Certificate | 4 (6) | 7 (9) | 11 (8) |
| Diploma | 9 (14) | 4 (5) | 13 (9) |
| Degree | 1 (2) | 0 (0) | 1 (1) |
| Patient diagnosis |  |  |  |
| Pneumonia | 35 (54) | 45 (61) | 80 (58) |
| Meningitis | 25 (38) | 28 (38) | 53 (38) |
| Malaria | 20 (31) | 14 (19) | 34 (24) |
| Gastroenteritis | 6 (9) | 14 (19) | 20 (14) |
| Malnutrition | 9 (14) | 5 (7) | 14 (10) |
| Bronchiolitis | 8 (12) | 5 (7) | 13 (9) |
| Anemia | 3 (5) | 6 (8) | 9 (6) |
| Sepsis/Septic Shock | 4 (6) | 4 (5) | 8 (6) |
| Dehydration | 3 (5) | 1 (1) | 4 (3) |
| Encephalitis | 0 (0) | 3 (4) | 3 (2) |
| Number of diagnoses |  |  |  |
| 1 | 22 (34) | 29 (39) | 51 (37) |
| 2 | 33 (51) | 25 (34) | 58 (42) |
| 3+ | 10 (15) | 20 (27) | 30 (22) |
| Case fatality |  |  |  |
| Case fatality for hospital duration | 7 (11) | 9 (12) | 16 (12) |
| Case fatality for initial 48hrs | 4 (57) | 4 (44) | 8 (50) |

Forty health care providers responded to the survey questions [supplemental material 2] reflecting on their experiences with the use of the FASTER monitoring tool, of which 14 (35%) were physicians, 22 (55%) nurses and 4 (10%) clinical officers (Table 4). Of 74 caregivers in the intervention arm, 70 (94%) responded to the survey questions (Table 5).

**Table 2:**
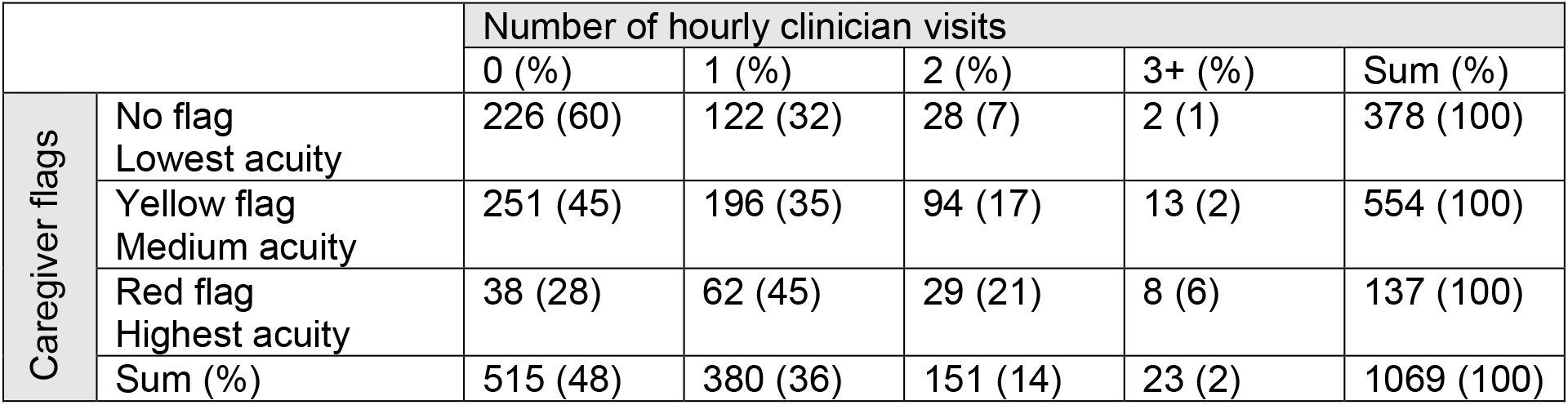
Number of hourly clinician visits by FASTER severity of illness level

**Table 3:** Number of clinician assessments per hour in intervention vs control group over the first 24hr enrollment period. Blank hourly caregiver data forms were counted as no clinician visits.

| Number of nurse and physician visits<br>at patient's bedside per hour | Control<br>n (%) | Intervention<br>n (%) |
| --- | --- | --- |
| 0 | 980 (65) | 1148 (67) |
| 1 | 384 (26) | 380 (22) |
| 2 | 116 (8) | 151 (9) |
| 3 | 15 (1) | 18 (1) |
| 4 | 5 (0) | 4 (0) |
| 5 | 1 (0) | 0 (0) |
| 7 | 0 (0) | 1 (0) |
| Number of physician visits at patient's<br>bedside per hour | Control<br>n (%) | Intervention<br>n (%) |
| 0 | 1246 (83) | 1406 (83) |
| 1 | 238 (16) | 276 (16) |
| 2 | 15 (1) | 19 (1) |
| 3 | 2 (0) | 1 (0) |
| Number of nurse visits at patient's<br>bedside per hour | Control<br>n (%) | Intervention<br>n (%) |
| 0 | 1118 (74) | 1284 (75) |
| 1 | 358 (24) | 399 (23) |
| 2 | 21 (1) | 16 (1) |
| 3 | 4 (0) | 2 (0) |
| 5 | 0 (0) | 1 (0) |

**Table 4:** Clinician survey of FASTER intervention

| Clinician | Physician | Nurse | Clinical officer | Sum |
| --- | --- | --- | --- | --- |
|  | Count (%) |  |  |  |
| Role | 14 (35) | 22 (55) | 4 (10) | 40 (100) |
| Days of exposure to FASTER intervention |  |  |  |  |
| 1-5 | 5 (36) | 8 (36) | 0 (0) | 13 (32) |
| 6-20 | 2 (14) | 8 (36) | 1 (25) | 11 (28) |
| >20 | 7 (50) | 6 (27) | 3 (75) | 16 (40) |
| Overall Impression of FASTER monitoring |  |  |  |  |
| Good | 8 (57) | 6 (27) | 2 (50) | 16 (40) |
| Not Good | 1 (7) | 6 (27) | 1 (25) | 8 (20) |
| Didn't notice much | 4 (29) | 6 (27) | 1 (25) | 11 (28) |
| Ambiguous/missing | 1 (7) | 4 (18) | 0 (0) | 5 (12) |
| Challenges of FASTER intervention: increased work |  |  |  |  |
| No | 10 (71) | 13 (59) | 3 (75) | 26 (65) |
| Yes | 4 (29) | 9 (41) | 1 (25) | 14 (35) |
| Challenges of FASTER intervention: false flags |  |  |  |  |
| No | 6 (43) | 16 (73) | 3 (75) | 25 (62) |
| Yes | 8 (57) | 6 (27) | 1 (25) | 15 (38) |
| Challenges of FASTER intervention: parents demanding |  |  |  |  |
| No | 8 (57) | 12 (55) | 3 (75) | 23 (57) |
| Yes | 6 (43) | 10 (45) | 1 (25) | 17 (42) |
| Would FASTER intervention improve care of a very sick child in resource-limited settings? |  |  |  |  |
| No | 1 (7) | 6 (27) | 0 (0) | 7 (18) |
| Yes | 13 (93) | 16 (73) | 4 (100) | 33 (82) |

**Table 5:** Caregiver feedback of FASTER intervention

| **Caregiver survey questions** | **Count (%)** |
| --- | --- |
| Number of caregivers (all female) | 70 (100) |
| Training easy to understand? |  |
| Yes | 63 (90) |
| No | 7 (10) |
| Clinicians responded as expected? |  |
| Yes | 51 (73) |
| No | 18 (26) |
| Challenge of FASTER intervention - fatigue |  |
| Yes | 13 (19) |
| No | 57 (81) |
| Challenge of FASTER intervention - stress |  |
| Yes | 18 (25) |
| No | 52 (75) |
| Challenges of FASTER intervention - clinician interaction |  |
| Yes | 3 (4) |
| No | 67 (96) |
| Challenges - other* |  |
| Yes | 8 (11) |
| No | 62 (89) |
| Would FASTER improve care very sick hospitalized child in this setting? |  |
| yes | 70 (100) |
| Would FASTER monitoring help recognize a sick child in your community? |  |
| yes | 70 (100) |

\*see qualitative data in results section

### Effectiveness of FASTER Monitoring Tool

On average, clinicians were significantly responsive to patient condition. Patients with admission PEWS of ≥ 10 received on average 0.79 (SD 0.89) and 0.70 (SD 0.40) visits/hour on the intervention and control arms, respectively, compared with only 0.39 (SD 0.20) and 0.34 (SD 0.14) with admission PEWS<5 (p<0.001 for linear association with PEWS). A similarly strong association was seen between hourly clinician visits and study-team FASTER scores (Table 2; Chi Squared p<0.001). Model estimates indicated that children in red-flag condition received 41% more visits on average than other children (p=0.002). However, there was no difference in provider responsiveness between the two arms (point estimate for the difference -0.2%, p=0.99) (Table 3). In other words, there was no observed intervention effect upon clinician behavior. In the same vein, examining whether there were clinical-course differences between the arms, among children with a 24-hour PEWS score the decrease from admission PEWS was not significantly different between the intervention and control arms (p=0.45).

There were 0.57 (SD 0.81) and 0.54 (0.76) visits/hour on average between 06:00 and 22:00 in the intervention and control arm, respectively. Nurse patient reassessments (0.32/hour on average during 06:00 – 22:00) were somewhat more frequent than physicians’ (0.24/hour).

There was a diurnal pattern in clinician patient interaction (Figure 2). Physicians’ visits peaked sharply around 09:00-10:00, with much fewer visits at other hours. Nurse visits peaked abruptly near 06:00, then retained a similar rate through most of the day, tapering off towards evening then dropping sharply late at night.

**Figure 2:**
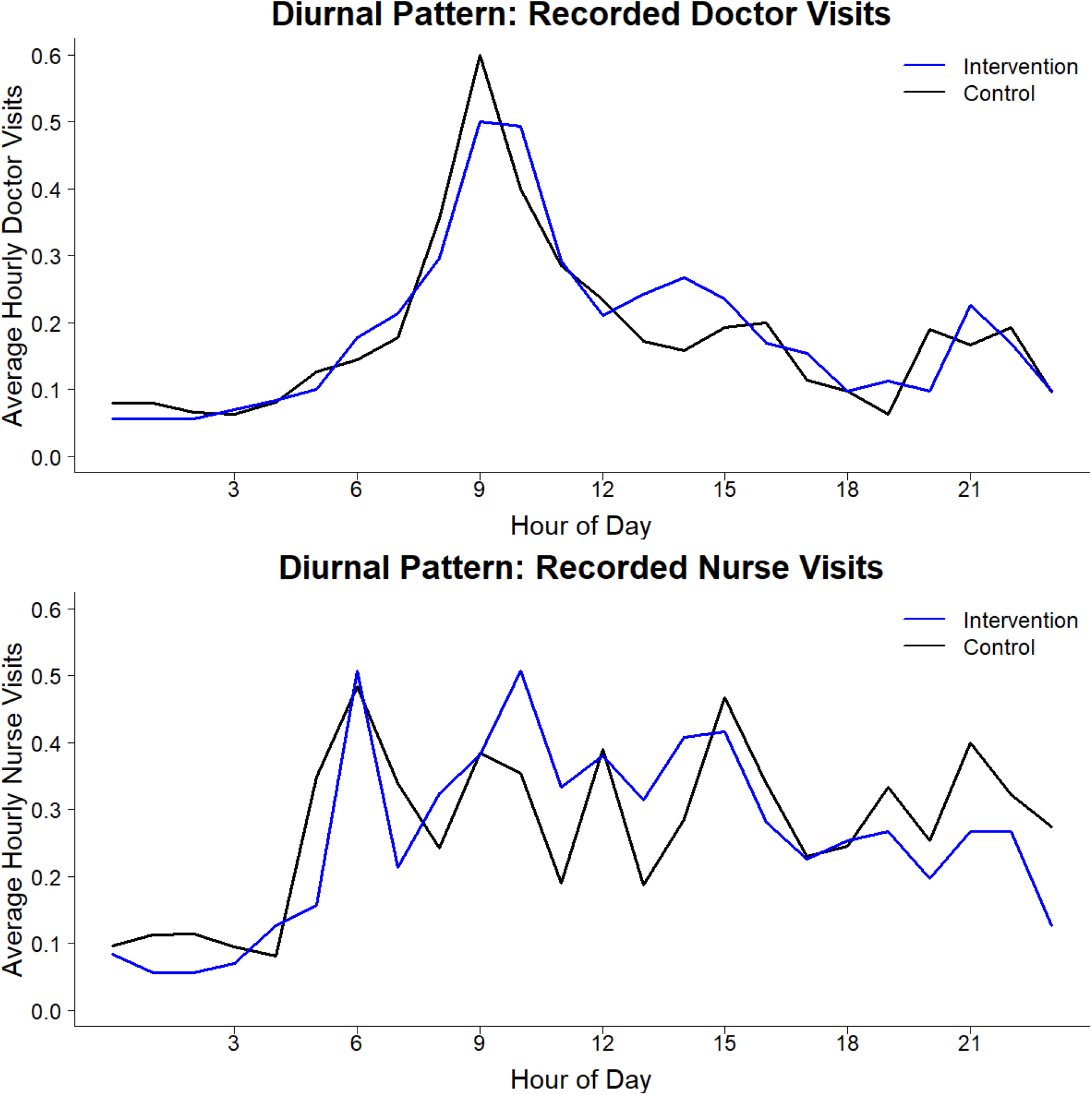
Daytime pattern of clinician patient visits in intervention versus control arm

### Overall Impression of FASTER Monitoring Tool

In response to an open-ended survey question, the overall impression of the FASTER monitoring tool was positive for 16 (40%) clinicians, not good for 8 (20%) and 16 (40%) did not notice a difference. Of those who reflected positively, the tool was described as an “innovative way for parents to get involved in the management process” and “an educative tool especially for parents who could identify danger signs”. Clinicians who rated FASTER negatively felt it did not work and was challenging. Of those who did not notice a difference, 9 (23%) reported that they did not see or rarely saw a flag, either indicating the caregivers were in the control arm or the children were doing well enough that no flags were raised.

### Feasibility of FASTER Monitoring Tool

Caregivers reported that the FASTER parental monitoring tool was easy to learn. Sixty-three (90%) caregivers reported that the FASTER training was easy to understand, whereas only seven caregivers (10%) described the FASTER training as “difficult” and “confusing” due to the complexity of monitoring, especially respiratory status, and raising flags. However, these caregivers also explained that with concentration and repetition, monitoring became easier.

Fifty-one (73%) caregivers felt clinicians responded appropriately to FASTER flags when raised. Only 18 (26%) reported clinicians did not respond as anticipated, either due to lack of enough clinicians or responses were delayed or not as frequent as expected.

The majority of caregivers (62 [88%]) felt supported and treated well by the clinicians who responded to their flags, whereas only a minority (8 [11%]) expressed that clinicians were not fully available or not kind.

In an open ended question, the majority of clinicians [26 (65%)] felt that parents could capture their child’s severity of illness and respond adequately all the time [23 (59%)] or sometimes [3(8%)]. A minority of clinicians [9 (23%)] was concerned that parents became too emotional and interpreted it [FASTER monitoring tool] as a “death sentence”. One clinician poignantly stated, “It [FASTER monitoring tool] is not a good idea, parents think we are killing their babies.”

### Challenges of FASTER Monitoring Tool

When asked with closed ended questions regarding the challenges of the tool, increased workload and false signaling, including “parents not pulling down flags,” were reported by 14 (35%) and 15 (38%) of clinicians respectively. Seventeen health care providers (42%) felt the monitoring tool was challenging to use because parents became more demanding. Through open ended questioning, another 17 clinicians reported additional challenges in using the FASTER tool, including 7 (18%) who explained that the monitoring intervention, especially the flag system, triggered very strong caregiver emotions, as the red flag “is a bad sign for their children” and “is like labeling a child very sick hence giving no hope”. They felt that the intervention may make caregivers “more anxious even when the child is not very sick.” A few clinicians also perceived the tool as too difficult to understand for caregivers (3 [8%]), and worried the flagging system, rather than a phone call, potentially delayed the response of clinicians (3 [8%]).

While the majority of caregivers (39 [56%]) reported that the monitoring intervention did not need any modifications or improvements, several caregivers provided suggestions, including: a) educate caregivers later in the admission once they are more “settled” (4 [6%]), b) change the frequency and timing of monitoring, as monitoring hourly at night is very difficult (2 [3%]), c) use a phone rather than flag to notify health care providers (1 [1%]), d) provide more education to caregivers on how to intervene if a red flag is raised (2 [3%]), and e) increase the monitoring performed by research staff or clinicians (3 [4%]).

Through open ended questioning, clinicians suggested improving the parental monitoring tool with additional education, training, and frequent reminders for both parents and clinicians (22 [55%]). Several clinicians (8 [20%]) also suggested using a different system than raising flags, given concerns about not seeing the flags in a timely manner, and that a red flag may represent a “bad omen”. They recommended using a bell or alarm system, especially when the hospital is busy. A few clinicians (5 [13%]) recommended increasing the number of hospital staff in general.

### Overall Value of FASTER Monitoring Tool in Resource Limited Settings

Despite challenges with the FASTER monitoring tool, the majority of clinicians [33 (82%)] agreed the FASTER intervention would improve care of a very sick child in a resource limited setting. When asked to explain their reasoning, some clinicians (11 [28%]) mentioned the monitoring tool increased the involvement of caregivers by improving their knowledge and ability to identify early warning signs. As one clinician commented, “It will allow mothers to raise their concerns and hence appropriate interventions where necessary leading to better outcome[s].”

Twenty (50%) clinicians felt the caregiver monitoring tool helped triage sicker patients first. One clinician stated, “It helps to signal the doctors that the patient/child needs urgent and quick medical attention which help[s] in early diagnosis and early management of the patient to save life.” Another explained, “When the flag is put [up] it helps us know the most sick child immediately and we act on it.” Seven (18%) clinicians found the parental monitoring tool especially helpful due to the scarcity of clinicians. As one clinician explained, “Since the health workers are limited, it [FASTER monitoring tool] would help in alerting where there is need.”

All caregivers (70 [100%]) agreed that FASTER monitoring would improve care of a very sick hospitalized child in their setting. Caregivers provided multiple open-ended explanations, including that with the increased knowledge, they could monitor the progress of their hospitalized child better, communicate better with health care providers, and alert medical staff earlier when the child was sicker or in “danger” and “might save the child’s life”. Similarly, 70 (100%) caregivers agreed their FASTER skills would help them recognize a sick child in their community. Through open-ended responses, several caregivers (14 [20%]) recommended broadening the scope of the intervention to outside the hospital settings and to more mothers, because the knowledge gained was so “helpful” and empowering. One mother explained, “Mothers will be empowered to act fast when the child is not doing well.” Another mother stated, “It enlightens you on how to be keen on monitoring your child. Even in the future it will still help me because I have learned.”

## Discussion

This study did not find a difference in the frequency of clinician visits to the patients’ bedsides between the FASTER intervention and control arms, nor was there a measurable health benefit in the study for patients receiving FASTER caregiver monitoring; although the study was not powered to detect the latter. Refinement of the implementation process of the FASTER tool is needed to improve its effectiveness particularly through greater acceptability and adoption by clinicians. However, the results of this pilot study add to the evidence (11) that the FASTER bedside assessment tool is feasible for caregivers of hospitalized children in low-resource settings and the tool was overall rated positively by both caregivers and clinicians.

As described by Lambert et al, early warning tools are more than just a ‘score’. They are part of a multifaceted ‘system’ approach to improve child patient safety and clinical outcomes(14).

Four integrated components are needed which work together to provide a comprehensive safety system for detection and management of the clinically deteriorating patient: (1) the afferent component which detects clinical deterioration and triggers an appropriate response such as the caregivers’ FASTER flag; (2) the efferent component consisting of the medical personnel providing the response, (3) the process improvement component containing elements such as auditing/monitoring/evaluation to enhance patient care and safety and (4) the governance/administrative component focusing on the organizational leadership, safety culture, education and processes required to implement and sustain the system (15). How these four components relate to the current and possible future FASTER implementation at KNH will be described here.

Our data suggest, both through non completion overnight and caregiver feedback, hourly monitoring, especially at night-time, is difficult and a monitoring schedule every 2-4 hours may be more feasible. In addition, FASTER flags were not always visible or noticed by clinicians and another form of alarm (bell versus phone) may be necessary to better trigger the response arm, as suggested by both caregivers and clinicians. Cultural concerns of red flags seen as bad omen need to be further explored with caregiver focus group discussions. Discussing death and prognosis has been described as a cultural taboo in Kenya given concerns of associated stigma and “inviting death”(16). Implementation of FASTER monitoring with sufficient caregiver education on goals to hasten interventions and without the label of a red flag may help address this issue. Given recent data on mortality predictions scores improving by including at least one element of the four top categories of altered consciousness, vital signs, signs of respiratory distress and indicators of malnutrition, addition of mid-upper arm circumference is important to consider to increase sensitivity and specificity of the FASTER tool (17).

The differences in caregiver versus clinician perceptions of the FASTER intervention may reflect the current paternalistic medical culture that is described at KNH and remains common in many parts of the world(18),(19). Through the FASTER monitoring, caregivers felt empowered and described a positive experience, whereas clinicians rated the intervention slightly less positively, describing one of the challenges of the tool as parents being “more demanding”. Since the study lacked resources for extensive outreach and preparation of clinicians, they may have experienced FASTER as a disruption or potential threat. While caregivers appreciated engaging constructively in the medical care of their children, clinicians were caught off-guard interacting with newly educated caregivers who felt empowered in assisting with clinical triage. Other “care by parent schemes” in which parents would assist with some nursing aspects of their hospitalized children (such as measuring temperature, giving medications) have described increased caregiver satisfaction and parents being capable of acceptable nursing care with little direction(20, 21). It is also possible, however, that caregivers did not feel comfortable sharing negative feedback regarding the monitoring tool as they shared their opinions through study nurses. Successful FASTER implementation may need to achieve improved “buy-in” from clinicians by emphasizing that the medical decision power remains with them, and that caregivers should be recognized as allies and assets, collecting data to help detect patient deterioration earlier so that medical interventions can be provided sooner. Hospital care with parental participation has previously been shown to help alleviate the workload of clinicians(22).

Despite resource limitations, clinicians focus their attention on the sickest children as indicated by the association between frequency of bedside visits and high Bedside PEWS and research team FASTER scores. Yet, given the observed 48 hour case fatality rate, much higher than in high resource settings, FASTER caregiver monitoring with modified implementation may enhance earlier recognition and management of clinical deterioration, especially at times with decreased clinician staffing. Based on suggestions from caregivers, expanding the educational aspects of the monitoring tool to mothers in the outpatient setting may also lead to earlier medical care seeking in the course of illness, leading to lower “early” fatalities.

The study was performed following a 100-day physician strike in Kenya and during a 151-day national nursing strike, in which the KNH nurses did not participate. Health care seeking behavior during the strike differed with pediatric patient volumes reduced by 20,000 compared to the prior year (23). Hence, the clinician response to the sickest patients may have been better during the study period as compared to the usual times with full volume pediatric wards. The diurnal pattern of physicians’ visits, with their presence focused between 09:00-10:00 for ward rounds and then diminished during the rest of the day is consistent with many physicians leaving the government hospital for their other sites of employment, reinforcing the importance of developing alternative methods to closely monitor patients in the afternoon, evening, and night hours.

Other factors in addition to hierarchical relationships and competing clinician priorities that have hindered the implementation of clinical best-practices at KNH in the past will need to be addressed in order to improve both the efferent clinician response as well as the process improvement for delivery of the FASTER intervention. Relevant factors include; 1) poor communication between nursing staff and physicians and central administration, 2) lack of objective mechanisms for monitoring and evaluating quality of clinical care due to inadequacies in clinicians’ self-regulation or motivation, 3) limited capacity for planning strategic change with chronic overcrowding of patients and staff being overworked, 4) limited management skills to introduce and manage change (18). Audit and feedback interventions with Kenyan pediatric health care providers and hospital administrators have, however, shown that they are committed to improving care, reinforcing quality standards, and enhancing team work (24). Utilizing different approaches that emphasize evidence in decision-making on innovation in healthcare might positively influence future FASTER implementation, e.g. with nurses in the acute sector shown to prefer a combination of practical (‘how to’) and scientific (‘principles’) knowledge, while medical professionals placing greater weight on the latter (25). Successful implementation of the FASTER tool in this complex environment will need to be more nuanced than simply training caregivers and clinicians. This will require working with focus groups of nursing, physician, and managerial stakeholders in addition to caregiver representatives to find culturally acceptable, effective, and sustainable ways to better integrate the FASTER tool into practice, achieve comprehensive buy-in and improve care.

There were several important limitations to this study. The study sample size was relatively small and limited to one site with a complex environment. Furthermore, the much lower prevalence of “red-flag” assessments meant that it was only powered to detect a very large intervention effect. The study was performed in a chronically strained healthcare system that had recently gone through further challenges following a prolonged physicians’ strike. The study occurred during Kenya’s presidential elections, during which political crises and violence led to medical and study staff intermittently not coming to work. Given the political situation and health care provider strikes, patient volumes were lower than usual. Hourly data collection by caregivers, especially at night was limited, likely secondary to caregiver fatigue and stress. This resulted in some missing data, including missing-data disparities between arms, making interpretation of results more difficult. Pediatric admission distribution rotating between four different wards led to decreased total exposure of the FASTER intervention per clinician and may have fostered unfamiliarity with the study and decreased recognition and response to caregiver flags. Clinician training was performed at the beginning of the study only, without auditing or performing further process improvements during the intervention period which may have contributed to decreased clinician participation in FASTER flag recognition.

Inpatient mortality remains unacceptably high in many low-income settings. The significant strains placed on limited numbers of clinicians suggest that interventions supporting the recognition of clinical deterioration may be beneficial. The FASTER tool appears to be feasible to implement and caregivers reported they felt empowered by the tool. In addition, caregivers felt supported by the additional education that was provided with the tool and requested that the scope of the intervention be expanded to outside the hospital setting. Additional studies of the FASTER tool following modifications to improve fidelity may improve effectiveness.

## Data Availability

All data requests would be subjected to approval of the IRBs cited above.

## Acknowledgements

This study was supported by the Seattle Children’s Research Institute’s Center for Clinical and Translational Research Academic Enrichment Fund. The content of this article is solely the responsibility of the authors and does not necessarily represent the official views of their respective employers or funders, who had no role in study design, data collection, data analysis, decision to publish, or preparation of the manuscript.

## References

1. UN IGME UN Inter-agency Group for Child Mortality Estimation. Child Mortality Estimates 2020. Available from: https://childmortality.org/. Accessed October 10, 2021

2. Global Burden of Disease Under-5-Mortality Collaborators. Global, regional, and national progress towards Sustainable Development Goal 3.2 for neonatal and child health: all-cause and cause-specific mortality findings from the Global Burden of Disease Study 2019. Lancet. 2021;398(10303):870–905.

3. World Health Organization. Health Workforce Requirements for Universal Health Coverage and the Sustainable Development Goals. Geneva, Switzerland; 2016.

4. Nolan T, Angos P, Cunha AJ, Muhe L, Qazi S, Simoes EA, et al. Quality of hospital care for seriously ill children in less-developed countries. Lancet. 2001;357(9250):106–10.

5. Bitwe R, Dramaix M, Hennart P. [Quality of care given to seriously ill children in a provincial hospital in central Africa]. Sante Publique. 2007;19(5):401–11.

6. Olson D, Davis NL, Milazi R, Lufesi N, Miller WC, Preidis GA, et al. Development of a severity of illness scoring system (inpatient triage, assessment and treatment) for resource-constrained hospitals in developing countries. Trop Med Int Health. 2013;18(7):871–8.

7. Murthy S, Adhikari NK. Global health care of the critically ill in low-resource settings. Ann Am Thorac Soc. 2013;10(5):509–13.

8. Cummings MJ, Goldberg E, Mwaka S, Kabajaasi O, Vittinghoff E, Cattamanchi A, et al. A complex intervention to improve implementation of World Health Organization guidelines for diagnosis of severe illness in low-income settings: a quasi-experimental study from Uganda. Implement Sci. 2017;12(1):126.

9. George EC, Walker AS, Kiguli S, Olupot-Olupot P, Opoka RO, Engoru C, et al. Predicting mortality in sick African children: the FEAST Paediatric Emergency Triage (PET) Score. BMC Med. 2015;13:174.

10. Muttalib F, Clavel V, Yaeger LH, Shah V, Adhikari NKJ. Performance of Pediatric Mortality Prediction Models in Low-and Middle-Income Countries: A Systematic Review and Meta-Analysis. J Pediatr. 2020;225:182–92 e2.

11. von Saint Andre-von Arnim AO, Kumar RK, Oron AP, Nguyen QP, Mutonga DM, Zimmerman J, et al. Feasibility of Family-Assisted Severity of Illness Monitoring for Hospitalized Children in Low-Income Settings. Pediatr Crit Care Med. 2021;22(2):e115–e24.

12. Parshuram CS, Duncan HP, Joffe AR, Farrell CA, Lacroix JR, Middaugh KL, et al. Multicentre validation of the bedside paediatric early warning system score: a severity of illness score to detect evolving critical illness in hospitalised children. Crit Care. 2011;15(4):R184.

13. Harris PA, Taylor R, Thielke R, Payne J, Gonzalez N, Conde JG. Research electronic data capture (REDCap)--a metadata-driven methodology and workflow process for providing translational research informatics support. J Biomed Inform. 2009;42(2):377–81.

14. Lambert V, Matthews A, MacDonell R, Fitzsimons J. Paediatric early warning systems for detecting and responding to clinical deterioration in children: a systematic review. BMJ Open. 2017;7(3):e014497.

15. Jagt EW. Improving Pediatric Survival from Resuscitation Events: The Role and Organization of Hospital-based Rapid Response Systems and Code Teams. Curr Pediatr Rev. 2013;9(2):158–74.

16. Love KR, Karin E, Morogo D, Toroitich F, Boit JM, Tarus A, et al. “To Speak of Death Is to Invite It”: Provider Perceptions of Palliative Care for Cardiovascular Patients in Western Kenya. J Pain Symptom Manage. 2020;60(4):717–24.

17. Ogero M, Sarguta RJ, Malla L, Aluvaala J, Agweyu A, English M, et al. Prognostic models for predicting in-hospital paediatric mortality in resource-limited countries: a systematic review. BMJ Open. 2020;10(10):e035045.

18. Irimu GW, Greene A, Gathara D, Kihara H, Maina C, Mbori-Ngacha D, et al. Factors influencing performance of health workers in the management of seriously sick children at a Kenyan tertiary hospital--participatory action research. BMC Health Serv Res. 2014;14:59.

19. Couet N, Desroches S, Robitaille H, Vaillancourt H, Leblanc A, Turcotte S, et al. Assessments of the extent to which health-care providers involve patients in decision making: a systematic review of studies using the OPTION instrument. Health Expect. 2015;18(4):542–61.

20. Sainsbury CP, Gray OP, Cleary J, Davies MM, Rowlandson PH. Care by parents of their children in hospital. Arch Dis Child. 1986;61(6):612–5.

21. Cleary J, Gray OP, Hall DJ, Rowlandson PH, Sainsbury CP, Davies MM. Parental involvement in the lives of children in hospital. Arch Dis Child. 1986;61(8):779–87.

22. Melo EM, Ferreira PL, Lima RA, Mello DF. The involvement of parents in the healthcare provided to hospitalzed children. Rev Lat Am Enfermagem. 2014;22(3):432–9.

23. Kaguthi GK, Nduba V, Adam MB. The impact of the nurses’, doctors’ and clinical officer strikes on mortality in four health facilities in Kenya. BMC Health Serv Res. 2020;20(1):469.

24. Wiaganjo P. Exploring the perceptions of pediatric health care workers on audit and performance feedback in Kenyan County Hospitals (Thesis). In: University S, editor. 2015.

25. Turner S, D’Lima D, Hudson E, Morris S, Sheringham J, Swart N, et al. Evidence use in decision-making on introducing innovations: a systematic scoping review with stakeholder feedback. Implementation Science. 2017;12.

